# Is acetazolamide a potential intervention for people with bipolar disorders? A scoping review protocol

**DOI:** 10.1101/2022.08.27.22279292

**Authors:** Rebecca Strawbridge, Stelios Orfanos, Nefize Yalin, Allan H Young

## Abstract

Acetazolamide is a carbonic anhydrase inhibitor whose primary indication is to alleviate altitude sickness, but it is also used in the treatment of glaucoma, seizures and oedema. Although it has not been repurposed for psychiatric illnesses, acetazolamide has been advocated for its putative benefits in those with a bipolar disorder (BD). Because current treatment options for BD bring a variety of challenges, it is important to examine the current evidence to date in a formal synthesis. This article describes a protocol for a scoping review to do so, in accordance with the Preferred Reporting Items for Systematic Reviews and Meta-Analyses extension for scoping reviews (PRISMA-ScR). PubMed, Embase and PsycInfo will be searched for all available publications focusing on the intervention acetazolamide for people with BD. This will include non-primary and primary research of any design, focusing on any outcome, in order to comprehensively bring together the work that has been undertaken to date on this topic. Relevant data from included articles will be subject to a narrative synthesis. It is anticipated that the results from the review will help to ascertain the potential for acetazolamide to benefit people with bipolar affective disorders in the future and aid the planning of future primary research to establish this.

## Introduction

Bipolar disorders (BD) are a leading cause of disability globally.^1^ Several factors contribute to this, including a high prevalence (∼2-4%),^2,3^ early onset and frequent lifetime episode recurrences,^4^ as well as the debilitating effects of mania, depression, functional and cognitive impairments which frequently persist in periods of remission from acute episodes, as well as subsyndromal symptoms between episodes.^4^ Notwithstanding the numerous effective treatments for people with BD, these frequently confer a variety of challenges ranging from risk of affective switch (to symptoms/episode of the opposite pole), tolerability, contraindication due to comorbidities or other medications taken, and variable interindividual clinical effectiveness.^5^ Some examples here are the safety profile of first-generation antipsychotics, negative symptoms and tolerability issues with second-generation antipsychotics, the unsuitability of valproate medications for women with childbearing potential, and the need for monitoring to ensure non-toxicity with lithium. These are acknowledged by well-regarded, widely used clinical guidelines^6,7^ and are supported by up-to-date systematic reviews and meta-analyses e.g., focused on acute treatment of mania,^8^ depression,^9^ maintenance treatment^10^ and rapid cycling.^11^ Each of the referenced syntheses here states a need for increased clinical trials, both of existing and new interventions.

Acetazolamide is a carbonic anhydrase inhibitor which reduces intraocular pressure and is used widely for a range of indications.12 It is currently licensed for glaucoma, epilepsy and abnormal fluid retention. Its anticonvulsant effects are in common with many therapies for BD. Since its first use as a diuretic ∼70 years ago, acetazolamide was first indicated for its putative value as a psychotropic almost 40 years ago, showing acute and prophylactic antipsychotic effects, particularly in patients with atypical and menstrual-exacerbated psychosis.^13^ Focus subsequently turned towards affective presentations.^14^ Because of the need for novel interventions, acetazolamide’s potential and an absence of evidence synthesis to date, we have developed a protocol for a scoping review to do so.

### Aims

To bring together all available evidence relating to the potential for acetazolamide to improve the lives of people with bipolar disorders. Given the limited size of the literature to date, we plan a broad and inclusive review not limited by research design or specific outcomes focused on. The specific questions we plan to answer are as follows, although the authors acknowledge that the review findings are largely dependent on the evidence available and thus are not limited to these questions.

1. Are there indications that acetazolamide can be effective for people with BD in terms of core affective symptomatology (mania, depression, mixed affective states)?
2. What are the putative effects of acetazolamide on broader important outcomes such as psychosocial and cognitive functioning and quality of life?
3. Are there indications that acetazolamide could be beneficial prognostically i.e., in reducing relapse?
4. How acceptable might acetazolamide be for patients to take in the short-and/or longer-term? This is considered in terms of adherence and continuation of the medication over time in combination with reports of tolerability and safety. We note that long-term use is generally cautioned with this intervention.
5. Are there indications of what might constitute a therapeutic dose and/or duration of acetazolamide for BD?

## Methods

### Design

This scoping review will adhere to the Preferred Reporting Items for Systematic Reviews and Meta-Analyses extension for scoping reviews (PRISMA-ScR) statement.^15^ The protocol for this review is being developed prior to the start of the review procedure to ensure transparency and rigor via pre-registration.

Following the development of specific research questions/scope (above) and publication of this protocol, an electronic database search will be undertaken. After duplicate records are removed, the review process consists of a) screening the titles and abstracts of all records; b) examining the full text of any potentially relevant article; c) extracting relevant data from any included article. These data will be narratively synthesised to answer the review questions to the greatest possible extent.

### Eligibility Criteria

To be included in the scoping review, articles must meet the following criteria.

A. Design; any (primary clinical or preclinical; non-primary).
B. Intervention: Acetazolamide (brand name: Diamox).
C. Participants (or population of interest/focus): Bipolar disorders.
D. Outcomes: any relevant to people with BD.

We will include both peer-reviewed articles, and any non-peer reviewed studies identified, with non-peer-reviewed studies weighted with less importance. We will include articles not written in English if it is feasible to obtain a sufficient translation. Where incomplete articles are retrieved, or in cases of missing relevant data, authors will be contacted with a request to provide additional necessary information.

### Search strategy

We will search PubMed, Embase and APA PsycInfo databases for all dates from inception until the search date. The following search terms will be used, searching all fields: *((Acetazolamide) or (Diamox)) AND ((bipolar) or (mania))*. Additionally, reference lists of included articles and a search of clinicaltrials.gov will be undertaken to bolster the database searches.

### Screening & data extraction procedure

Duplicate records retrieved will initially be de-duplicated using Ovid’s built-in tool, and subsequently in Rayyan open-source review management software.^16^ All articles will be imported into Rayyan, which will record the status of each article during the screening and inclusion process. One author (RS, SO or NY) will screen the title and abstracts for an initial indication of eligibility; subsequently, the full text of any potentially-eligible article will be examined in full. Any uncertainties will be resolved via discussion and consensus with one other reviewer, and subsequently all included articles will be agreed by the full review team.

Relevant data from all included articles will be extracted by one author (RS, SO or NY) using a standardised form (using Excel software). This information will include bibliography (e.g., authorship, publication), methodological characteristics (design, location, objectives, participants, interventions, outcomes) and results (see below). Any uncertainties will be resolved as described above. No formal methodological quality or risk of bias assessment will be undertaken, as standard for scoping reviews, however methodological factors will be considered and incorporated into the results and discussion.

### Outcome synthesis

In accordance with the review aims and questions stated above, we will focus on any outcome relevant to people with bipolar disorder, primarily those related to efficacy and acceptability/tolerability. Information relevant to future clinical trial planning will be considered important. Methodological/bibliographic characteristics will be summarised using tables and text. Results will be synthesised narratively, under categories as specified in the review questions section above. The categories/outcomes eventually used will vary depending on data availability. The narrative synthesis will explore patterns/themes arising from included articles, incorporating simple statistical data (e.g. report or calculate, where feasible, standardised effect sizes and measures of variance, or simpler descriptive data), and trend observations.

## Data Availability

Data produced in this study will be available upon reasonable request to the authors.

## Data availability

Data produced in this study will be available upon reasonable request to the authors.

## References

1. World Health Organization: The global burden of disease:… - Google Scholar [Internet]. [cited 2021 Sep 4]. Available from: https://scholar.google.com/scholar_lookup?title=The+global+burden+of+disease:+2004+update…publication_year=2008&

2. Merikangas KR, Akiskal HS, Angst J, Greenberg PE, Hirschfeld RMA, Petukhova M, et al. Lifetime and 12-Month Prevalence of Bipolar Spectrum Disorder in the National Comorbidity Survey Replication. Arch Gen Psychiatry. 2007 May 1;64(5):543–52.

3. Strawbridge R, Alexander L, Richardson T, Young AH, Cleare AJ. Is there a ‘bipolar iceberg’ in UK primary care psychological therapy services? Psychol Med. 2022 Aug 3;1–10.

4. Vieta E, Langosch JM, Figueira ML, Souery D, Blasco-Colmenares E, Medina E, et al. Clinical management and burden of bipolar disorder: results from a multinational longitudinal study (WAVE-bd). Int J Neuropsychopharmacol. 2013 Sep 1;16(8):1719–32.

5. Bauer M, Andreassen OA, Geddes JR, Vedel Kessing L, Lewitzka U, Schulze TG, et al. Areas of uncertainties and unmet needs in bipolar disorders: clinical and research perspectives. Lancet Psychiatry. 2018 Nov 1;5(11):930–9.

6. Goodwin GM, Consensus Group of the British Association for Psychopharmacology. Evidence-based guidelines for treating bipolar disorder: revised second edition--recommendations from the British Association for Psychopharmacology. J Psychopharmacol Oxf Engl. 2009 Jun;23(4):346–88.

7. Yatham LN, Kennedy SH, Parikh SV, Schaffer A, Bond DJ, Frey BN, et al. Canadian Network for Mood and Anxiety Treatments (CANMAT) and International Society for Bipolar Disorders (ISBD) 2018 guidelines for the management of patients with bipolar disorder. Bipolar Disord. 2018 Mar;20(2):97–170.

8. Yalin N, Strawbridge R, Dawson K, Fouani A, Young A. Monotherapy versus combined therapies for acute mania: a systematic review of benefits and harms [Internet]. [cited 2022 Aug 25]. Available from: https://www.crd.york.ac.uk/prospero/display_record.php?RecordID=309749

9. Jauhar S, Tong C, Strawbridge R, Arnone D, Young A. Efficacy of therapeutic interventions in bipolar depression: a systematic review and network meta-analysis [Internet]. [cited 2022 Aug 25]. Available from: https://www.crd.york.ac.uk/prospero/display_record.php?RecordID=227477

10. Nestsiarovich A, Gaudiot CES, Baldessarini RJ, Vieta E, Zhu Y, Tohen M. Preventing new episodes of bipolar disorder in adults: Systematic review and meta-analysis of randomized controlled trials. Eur Neuropsychopharmacol. 2022 Jan 1;54:75–89.

11. Strawbridge R, Kurana S, Kerr-Gaffney J, Jauhar S, Kaufman KR, Yalin N, et al. A systematic review and meta-analysis of treatments for rapid cycling bipolar disorder. Acta Psychiatr Scand [Internet]. [cited 2022 Aug 25];n/a(n/a). Available from: https://onlinelibrary.wiley.com/doi/abs/10.1111/acps.13471

12. Farzam K, Abdullah M. Acetazolamide. In: StatPearls [Internet]. Treasure Island (FL): StatPearls Publishing; 2022 [cited 2022 Aug 25]. Available from: http://www.ncbi.nlm.nih.gov/books/NBK532282/

13. Inoue H, Hazama H, Hamazoe K, Ichikawa M, Omura F, Fukuma E, et al. Antipsychotic and prophylactic effects of acetazolamide (Diamox) on atypical psychosis. Folia Psychiatr Neurol Jpn. 1984;38(4):425–36.

14. Hayes SG. Acetazolamide in Bipolar Affective Disorders. Ann Clin Psychiatry. 1994 Jan 1;6(2):91–8.

15. Tricco AC, Lillie E, Zarin W, O’Brien KK, Colquhoun H, Levac D, et al. PRISMA Extension for Scoping Reviews (PRISMA-ScR): Checklist and Explanation. Ann Intern Med. 2018 Oct 2;169(7):467–73.

16. Ouzzani M, Hammady H, Fedorowicz Z, Elmagarmid A. Rayyan—a web and mobile app for systematic reviews. Syst Rev. 2016 Dec 5;5(1):210.

